# Beyond ICD Codes: Fine-Tuning LLMs for In-Hospital Cardiac Arrest Identification from EHR Notes

**DOI:** 10.64898/2026.09.24.26363973

**Authors:** Davy Weissenbacher, Jonathan Vo, Audrey Uy-Evanado, Kyndaron Reinier, Harpriya Chugh, Vishnu Kadiyala, Karen O’Connor, Sumeet S. Chugh, Graciela Gonzalez-Hernandez

## Abstract

**Background:** Identification of in-hospital cardiac arrest (IHCA) through manual chart abstraction is the gold standard, but its time-consuming and resource-intensive, which naturally limits its practicality. Diagnostic codes provide an accessible automated alternative, but prior work has shown this method of extraction has poor sensitivity and poor positive predictive value. More accurate and scalable automated methods are needed to support IHCA surveillance and research.

**Methods:** We developed large language model (LLM)-based systems that incorporated physician-defined clinical knowledge through guideline-enriched prompting, supervised finetuning, and a self-correction procedure. The systems identified IHCA events and their locations from electronic health record notes. Performance was evaluated at the encounter level against physician-adjudicated chart review and compared with ICD-code–based identification.

**Results:** Compared with an ICD-code–based algorithm (precision 0.68, recall 0.91, F1 score 0.78), the supervised fine-tuned LLM offered substantially better performance for IHCA identification on a curated evaluation corpus, reaching a precision of 0.88, a recall of 1.00, and an F1 score of 0.94. When tested on a larger, unselected validation cohort of 45,525 encounters, a separate LLM configuration—relying on guideline-enriched prompting and a self-correction procedure rather than fine-tuning—achieved a precision of 0.79 for encounter-level IHCA identification, meaning most encounters flagged by the model were genuine IHCA cases. The fine-tuned LLM underperformed on this cohort due to its inability to self-correct its decisions, an issue that can be mitigated by having a separate model perform the correction step.

**Conclusion:** Explicit integration of clinical knowledge through prompting and supervised fine-tuning enabled accurate identification of IHCA events and locations from routine clinical documentation. In a two-pass workflow, these systems could prioritize encounters for clinician adjudication, reduce manual screening requirements, and support more scalable IHCA surveillance and registry development.

## 1. Introduction

In-hospital cardiac arrest (IHCA) affects approximately 300,000 patients annually in the United States, corresponding to 1.6 to 2.8 events per 1000 hospital admissions.^1^ Survival to hospital discharge remains no higher than approximately 25%, underscoring the substantial mortality associated with these events.^2^ IHCA is conventionally defined as cessation of cardiac activity in a hospitalized patient requiring chest compressions and/or defibrillation as part of cardiopulmonary resuscitation (CPR).^3^

Despite advances in resuscitation care, the incidence of IHCA has continued to increase, while improvements in survival have recently plateaued.^1,4^ These trends highlight the need for accurate surveillance systems that can support benchmarking, outcomes research, and data-driven quality improvement. Accordingly, the American Heart Association’s “Ten Steps Toward Improving In-Hospital Cardiac Arrest Quality of Care and Outcomes” identifies the development of high-quality IHCA registries as a central component of efforts to improve resuscitation care.^5^

Current IHCA research relies heavily on prospective registries in which trained personnel manually abstract information from medical records and cardiac arrest flow sheets. The largest such registry in the United States is the American Heart Association’s *Get With The Guidelines–Resuscitation* registry.^6^ However, only 5–6% of US hospitals participate in the registry in a given year, limiting the representativeness and reach of registry-based surveillance.^7^ Expanding participation will require methods that preserve the clinical validity of expert adjudication while reducing the associated abstraction burden.

Outside prospective registries, IHCA cohorts are generally identified through manual chart review or through International Classification of Diseases (ICD) diagnosis and procedure codes extracted from the electronic health record (EHR).^8,9^ Manual review provides clinically detailed case ascertainment but requires substantial time and specialized expertise. ICD-based methods are considerably more scalable, but they have demonstrated limited sensitivity and positive predictive value for IHCA because of missing codes, miscoding, and codes carried forward from previous encounters.^8,9^ Regional and temporal variation in coding practices further limits their reliability for longitudinal surveillance and interinstitutional benchmarking.^10^

Clinical notes provide a richer representation of the events, interventions, and contextual information needed to determine whether an IHCA occurred. Natural language processing (NLP) and large language models (LLMs) have been applied to clinical text to identify cardiovascular phenotypes and risk factors, including heart failure, recurrent atrial fibrillation, and heart disease risk factors.^11–14^ To our knowledge, however, prior work has not addressed automated IHCA case identification from unfiltered clinical documentation. This task requires more than recognition of cardiac-arrest terminology: a system must apply clinically defined criteria, distinguish current events from historical or hypothetical mentions, determine whether resuscitative treatment occurred, and resolve whether the event began in an inpatient unit, the emergency department, or outside the hospital.

In this study, we investigate two complementary approaches for integrating physician-defined IHCA knowledge into LLMs. First, we encode clinical definitions and annotation rules explicitly through guideline-enriched prompting, few-shot examples, and a self-correction procedure. Second, we incorporate the same clinically adjudicated knowledge into model parameters through supervised fine-tuning. We develop and evaluate LLM-based systems for identifying IHCA events and their locations from unfiltered EHR notes, using physician-adjudicated chart abstraction as the reference standard and ICD-code–based identification as the principal comparator.

## 2. Materials and Methods

### 2.1. Data Compilation and Annotation

The study was approved by the Cedars-Sinai Medical Center Institutional Review Board. We constructed two datasets for model development and evaluation. The first, termed the *Evaluation Corpus*, comprised 500 clinical encounters, defined as a complete episode of care from intake through discharge, from the Cedars-Sinai Medical Center electronic health record (EHR). An individual patient could contribute more than one encounter. For each encounter, the corpus included the available inpatient documentation, including emergency department (ED) notes, discharge summaries, and Code Blue notes recorded following cardiac arrest resuscitation. An encounter could contain multiple notes of the same type.

To obtain sufficient positive examples for model development, the Evaluation Corpus combined code-enriched and randomly sampled encounters. Of the 500 encounters, 47% contained at least one diagnosis or procedure code selected to identify a potential cardiac arrest, while the remaining 53% were randomly sampled encounters. Code presence was used only for cohort enrichment and was not treated as evidence that an IHCA had occurred. The codes used for sampling are listed in Table 1.

**Table 1.** Diagnosis and procedure codes used to enrich the Evaluation Corpus for potential cardiac arrest encounters.

| Code | Description |
| --- | --- |
| I49.01 | Ventricular fibrillation |
| I49.02 | Ventricular flutter |
| I46.2 | Cardiac arrest due to underlying cardiac condition |
| I46.8 | Cardiac arrest due to other underlying condition |
| I46.9 | Cardiac arrest, cause unspecified |
| 92950 | Cardiopulmonary resuscitation |
| 5A12012 | Performance of cardiac output, single, manual |
| 02QA0ZZ | Repair heart, open approach |

Before annotation began, the two lead investigators iteratively developed annotation guidelines grounded in the Utstein definition of IHCA as pulselessness requiring chest compressions and/or defibrillation in a hospitalized patient.^3^ Two physicians (JV and AUE) independently applied the draft guidelines to a pilot sample of 50 notes. They adjudicated disagreements and refined the guidelines until no further modifications were required. The final annotation guidelines were previously published by Vo et al.^15^

Using these guidelines, JV and AUE annotated 3,822 notes from the 500 encounters. The physician annotations served as the reference standard. A note was labeled *IHCA* when the clinical text clearly documented a pulseless event treated with chest compressions and/or external defibrillation; otherwise, it was labeled *not IHCA*. Labels were based on whether resuscitative treatment was delivered rather than on whether treatment was clinically indicated or concordant with the patient’s code status. Thus, a pulseless event was labeled *IHCA* when chest compressions and/or external defibrillation were performed, including when the patient had a pre-existing do-not-resuscitate or comfort-care order. A pulseless event for which resuscitation was not attempted was labeled *not IHCA*.

Identifying all episodes of in-hospital pulselessness, including events without attempted resuscitation, represents an important but distinct surveillance objective relevant to end-of-life care and adherence to treatment limitations. Such events were outside the scope of this study, which was designed to align with Utstein-style IHCA registries focused on cardiac arrests for which resuscitation was attempted.^3^

For each note labeled positive for IHCA, we also annotated the location of the arrest. Location was categorized as: 1) in-hospital (IH), including critical care units, operating rooms, and cardiac catheterization laboratories; 2) emergency department (ED); and 3) out-of-hospital cardiac arrest for which treatment was initiated out of the hospital and continued on arrival to the emergency department (OH-ED). The third category, OH-ED (out-of-hospital cardiac arrest with continued treatment in the ED), is not conventionally included in standard IHCA definitions; its inclusion here was intended to facilitate identification of this subgroup and permit its exclusion in future research, as appropriate. For notes documenting multiple arrests, only the first arrest was labeled.

Following annotation of each individual note within an encounter, two physicians reviewed all notes for each encounter to adjudicate IHCA status and location at the encounter level. If there were multiple IHCA events in one encounter, the encounter-level location was adjudicated from the first event.

We randomly split the Evaluation Corpus into a training set (300 encounters, 60%), development set (50 encounters, 10%), and test set (150 encounters, 30%). The training set included 99 encounters with at least one IHCA event (including a total of 172 notes labeled positive for IHCA). The development set contained 11 positive encounters (including 22 IHCA positive notes), and the test set 37 positive encounters (with 70 IHCA positive notes). Two physicians independently annotated the same 50 encounters for presence and location of IHCA, noting a Cohen’s kappa of 0.951 for IHCA presence (almost perfect agreement) and 0.838 for IHCA location (strong agreement).^16^

Because the Evaluation Corpus was enriched with potential IHCA cases identified using diagnosis and procedure codes, its IHCA prevalence was higher than that expected in routine hospital data. We therefore conducted an additional evaluation using a year of encounters from the ongoing Observational Study of Sudden Cardiac Arrest (OSCAR) cohort.^17^ (dated between January 1 and December 31, 2023 (*n* = 45,525)), including inpatient, emergency department, and observation encounters, as well as encounters containing a Code Blue note. Ambulatory encounters were excluded. No additional code-based enrichment or case preselection was applied.

The LLM was applied to all encounters in the 2023 OSCAR subset, and encounters predicted as IHCA were reviewed by two physicians (JV and AUE).^17^ For each model-flagged encounter, the physicians reviewed all flagged notes and determined whether the event met the Utstein definition of IHCA.^3,17^ Encounters meeting this definition were classified as true positives, whereas the remaining model-flagged encounters were classified as false positives. Because model-negative encounters were not systematically adjudicated, this analysis was designed to estimate precision rather than recall or overall classification accuracy.

### 2.2. LLM choice and prompt development

As an initial step, we developed a strong off-the-shelf, prompt-based system to establish a performance baseline before supervised fine-tuning. We selected Meta’s LLaMA 3.1-405B instruction-tuned model because it is an open-weight model available with strong general instruction-following capabilities that can be deployed within on-premises computing environment, although it requires substantial computational resources.^18^

We evaluated different approaches, beginning with two prompt-based baselines. In the first baseline approach (line 2 in Table 2), the LLM was given a role description, a concise statement of the task (classifying notes mentioning IHCA and location), and the note text. In the second baseline (line 3 in Table 2), we augmented this prompt with a summary of the clinical guidelines used for annotation, concise definitions of clinical concepts, and rule-based restatements of the guidelines to compensate for the model’s limited exposure to clinician-authored medical documents. We then enhanced this second baseline using few-shot and chain-of-thought prompting.^19^ Few-shot prompting was implemented by adding a 10 training examples with correct IHCA and location labels, selected through iterative error analysis to highlight common false positives as well as representative true positives and true negatives. Chain-of-thought prompting, asking the LLM to briefly justify its predictions and cite supporting phrases, aka quotes, from the note, promotes step-by-step reasoning to improve explainability and encourage grounding.^19^ Finally (line 4 in Table 2) added a self-correction step on top of this enhanced prompt. For each note, we first obtained the model’s classification and reasoning, then triggered a second review only for notes initially classified as IHCA, as most remaining errors involved false positives or misassigned locations. In this review, the model received the initial classification, location, reasoning, and note text, and was instructed to confirm or revise its decision using one of three location-specific prompts (IH, ED, or OH-ED).

**Table 2.** IHCA detection performance at the Encounter-level: Evaluation and OSCAR 2023 Corpora. SC stands for self-correction, FS for few-shots, SFT for supervised fine-tuning.

|  | Evaluation corpus |  |  | OSCAR 2023 corpus |
| --- | --- | --- | --- | --- |
|  | Prec. | Rec. | F1 | Prec. |
| 1. ICD Codes | 0.68 | 0.91 | 0.78 | — |
| 2. Llama 405B - Basic Prompt | 0.77 | **1.0** | 0.87 | — |
| 3. Llama 405B - Final Prompt + FS | 0.86 | **1.0** | 0.93 | 0.47 |
| 4. Llama 405B - Final Prompt + FS + SC | **0.88** | 0.97 | 0.92 | **0.79** |
| 5. Qwen 3 4B - Final Prompt + FS + SC | 0.86 | 0.73 | 0.79 | — |
| 6. Qwen 3 4B - Final Prompt + SFT + SC | **0.88** | **1.00** | **0.94** | 0.59 |

### 2.3. Supervised full fine-tuning LLM

In the second step of our approach, we investigated training the model to further improve performance. We adopted a straightforward approach: fully fine-tuning its weights with supervision on labeled examples representative of the task at hand, leaving more complex alternatives to future work due to time constraints. With numerous open-source LLMs now available spanning different sizes and performance levels, we chose the Qwen 3 series^20^ for our experiments. This series includes both dense and mixture-of-experts models ranging from 0.6 to 235 billion parameters and is specifically optimized for reasoning. Computational and time constraints led us to select the smaller Qwen3-4B Instruct variant. We conducted fine-tuning using the Transformer Reinforcement Learning (TRL) library,^21^ chosen for its integration with Hugging Face Transformers.

To train the model, we presented each note and, using the same final prompt as for the system in line 3 of Table 2, asked the model to generate a JSON object indicating whether the note mentioned an in-hospital cardiac arrest and, if so, where it occurred (IH, ED, or OH-ED), along with supporting quotes from the text and a short explanation for its decision. We corrected the model’s output whenever it diverged from labels assigned by our physicians; for notes labeled as mentioning an in-hospital cardiac arrest, we provided a single standardized explanation—”The patient experienced a cardiac arrest and received treatment for it”—along with the supporting quotes as the correction. We trained the model for 8 epochs, optimizing the library’s default token-level cross-entropy loss function, and selected the last checkpoint. We compared the performance of this model against a simple baseline (line 5): the out-of-the-box Qwen3-4B Instruct model, prompted with our final prompt and our 10 few-shot examples.

### 2.4. Testing and validation

We evaluated the performance of each baseline and the final system on the test set of the Evaluation Corpus at the encounter level. Encounter-level IHCA predictions were chosen over note-level predictions because clinical and research applications depend on whether a patient experienced IHCA during an encounter, rather than whether an individual note mentions IHCA, and because ICD codes also identify IHCA at the encounter level.

We also compared LLMs performance to performance of ICD codes commonly used to identify IHCA (ICD codes I49.01, I49.02, I46.2, I46.8, I46.9, CPT code 92950, and procedure codes 5A12012 and 02QA0ZZ). ICD performance was evaluated in two steps. To assess recall, these codes were applied to a gold-standard list of known IHCA encounters from the annotated corpus, and any positive encounters not captured by the codes were counted as false negatives. To assess precision, we applied the same codes to encounters from the larger OSCAR study between 7/1/2021 and 10/31/2021 (2021 OSCAR subset), and a physician adjudicated each candidate as IHCA or not; encounters adjudicated as not IHCA were counted as false positives. We summarized binary encounter-level performance for IHCA detection using precision, recall, and F1 score: precision (equivalent to positive predictive value) denotes the proportion of predicted IHCAs that are true IHCAs, recall (equivalent to sensitivity) denotes the proportion of true IHCAs correctly identified, and F1 score denotes the harmonic mean of the two.

Multiclass performance for IHCA location was assessed using a confusion matrix, with correct predictions appearing on the diagonal (see Figure 1).^22^

**Fig. 1:**
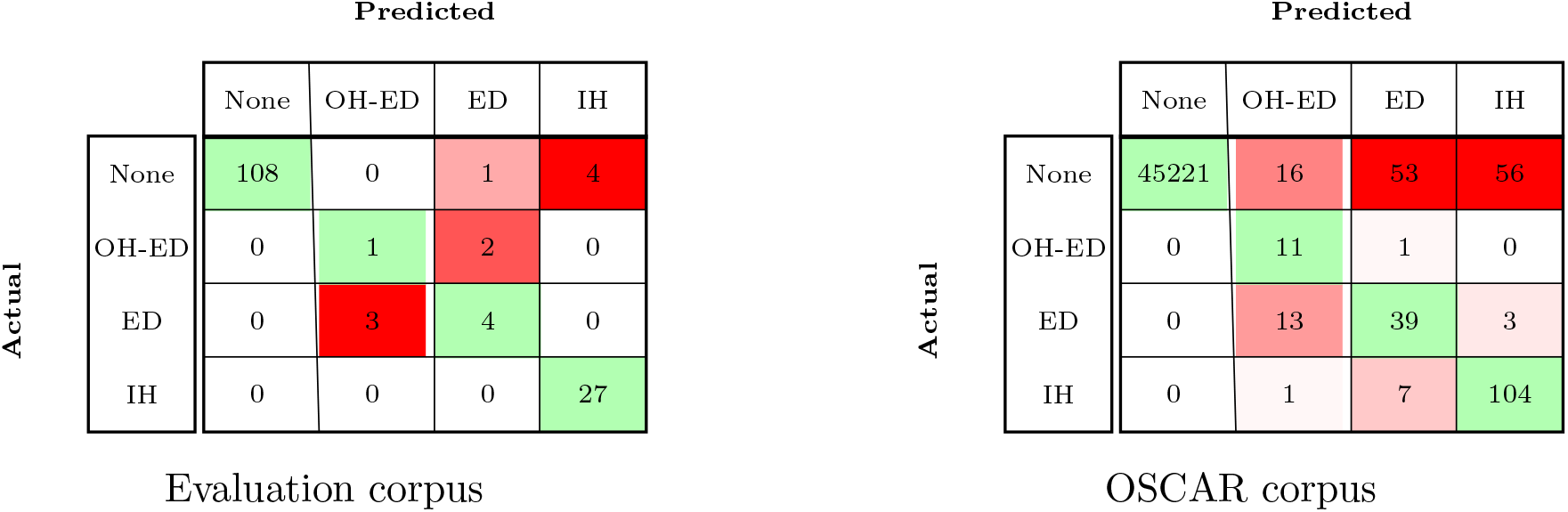
Confusion matrices for encounter-level IHCA prediction by the fine-tuned Qwen3-4B model on the test set of the Evaluation Corpus (left) and the OSCAR Corpus (right). Values along the diagonal indicate correct predictions; values in red indicate incorrect predictions. None refers to no IHCA. OH-ED refers to SCA that begins outside of the hospital with treatment continuing in the ED. ED refers to arrest that begins in the ED. IH refers to arrest that begins while admitted to the hospital.

## 3. Results

Table 2 details the performance of all systems. First, all generative systems achieve higher F1 scores than the ICD-based system (line 1) on the test set of the Evaluation corpus, regardless of the settings used to generate answers. Second, our prompt refinement produced a 6-point F1 gain between the systems in line 2 and line 3, on the test set of the Evaluation corpus. Adding the self-correction step then reduced recall from 1.0 to 0.97 (line 3 vs. line 4), while precision increased from 0.86 to 0.88 on the Evaluation corpus and from 0.47 to 0.79 on the OSCAR corpus. Finally, full fine-tuning supervision produced the largest F1 gains: the best-performing system (line 6) achieved perfect recall on the Evaluation corpus test set and an F1 score of 0.94, compared to 0.92 for LLaMA-405B, a model 100 times larger. On the OSCAR corpus, precision for this system achieve a low 0.59 compared to 0.79 for for LLaMA-405B. Note that, on the Evaluation corpus, although the fine-tuned Qwen3-4B model (line 6) outperformed LLaMA-405B (line 4), the difference did not reach statistical significance (paired bootstrap on binary note-level classification: 95% CI [-0.002, 0.097], *p* = 0.068). Despite a substantial overall sample size (N=1203), the relatively small number of positive cases (n=70) limited statistical power to detect differences in F1 score between systems. By contrast, on the OSCAR corpus, the LLaMA-405B (line 4) achieved significantly higher precision than the fine-tuned Qwen3-4B model (line 6): (paired bootstrap on binary note-level classification, restricted to notes flagged by at least one system: 95% CI [-0.142, -0.067], *p <* 0.001).

## 4. Discussion

This study demonstrates that physician-defined clinical knowledge can be incorporated into LLM-based IHCA identification through both contextual and parametric adaptation. Guideline-enriched prompting, few-shot examples, and targeted self-correction substantially improved the performance of an off-the-shelf LLaMA 3.1-405B model, while supervised full fine-tuning enabled a much smaller Qwen3-4B model to achieve the best overall performance. On the Evaluation Corpus, the fine-tuned Qwen3-4B model achieved a precision of 0.88, recall of 1.00, and F1 score of 0.94. These findings extend our previous work on LLM-based IHCA identification by showing that task-specific supervised adaptation can encode clinically adjudicated definitions and decision rules within a smaller model.^15^

The progression across prompt configurations also illustrates the value of explicitly representing clinical knowledge. The basic prompt required the model to classify a note using only a role description and task statement. Adding the physician-developed annotation guidelines, clinical definitions, selected examples, and evidence-grounded rationale instructions increased F1 from 0.87 to 0.93. The self-correction step produced a modest reduction in recall but improved precision, particularly in the lower-prevalence OSCAR cohort. These results suggest that explicit decision criteria and targeted review of likely error cases can improve discrimination between true IHCA events and clinically similar documentation, including historical arrest mentions, ED arrests, and out-of-hospital arrests with continued treatment in the ED. Supervised fine-tuning produced an additional improvement over prompting alone. The fine-tuned Qwen3-4B model achieved an F1 score of 0.94, compared with 0.92 for the prompted LLaMA 3.1-405B model with self-correction. Although this difference did not reach statistical significance on the Evaluation Corpus, the finding is operationally relevant: a 4-billion-parameter model may be more feasible for secure local deployment than a 405-billion-parameter model. The result also indicates that task-specific clinical supervision can partially compensate for differences in model scale.

The OSCAR analysis addressed a different question from the curated Evaluation Corpus. Rather than estimating complete classification performance, it evaluated the precision of model-positive predictions in a year-long cohort without code-based enrichment. The prompted LLaMA model’s precision increased from 0.47 to 0.79 after self-correction, indicating that the second-pass review substantially reduced false-positive screening results. The final precision of the fine-tuned Qwen3-4B model was lower with 0.59. Because model-negative OSCAR encounters were not systematically adjudicated, recall, overall accuracy, and the underlying IHCA prevalence cannot be estimated from this analysis. Accordingly, the OSCAR findings support the model’s potential utility for prioritizing encounters for review but do not constitute external validation or a complete population-level performance assessment.

These findings have practical implications for IHCA surveillance. Manual abstraction remains necessary for high-validity registry construction but requires substantial clinical effort.^2^ A two-stage workflow could use the model to screen routine documentation and present likely IHCA encounters, together with supporting text, for clinician adjudication. Such an approach could reduce the number of records requiring full review while preserving human oversight. However, the present study did not directly measure abstraction time, number needed to review, reviewer agreement with model-provided evidence, computational cost, or workflow integration. These operational outcomes should be evaluated prospectively before claims of reduced workload or improved registry participation can be confirmed. If validated across institutions, such systems could lower barriers to participation in IHCA surveillance and quality-improvement initiatives.^5^

The study also highlights limitations in the clinical knowledge available to general-purpose LLMs. Publicly available models are trained predominantly on nonclinical text, and their performance may be constrained by limited exposure to clinical documentation and institution-specific language.^23,24^ Guideline-enriched prompting partially addresses this limitation by supplying relevant knowledge at inference time, while supervised fine-tuning incorporates task-specific knowledge into model parameters. Further adaptation using clinical documents and reinforcement learning with human-feedback may improve robustness across note types and documentation styles.^25,26^

Several limitations warrant emphasis. First, all data were obtained from a single health system. Documentation conventions, Code Blue workflows, note templates, and terminology may differ across institutions and EHR platforms. Evaluation at independent health systems is therefore necessary before generalizability can be established.

Second, the Evaluation Corpus was enriched with encounters containing cardiac arrest-related codes and included relatively few positive encounters in the test set. Performance estimates, particularly the observed recall of 1.00, should therefore be interpreted with appropriate uncertainty.

Third, only model-positive encounters were adjudicated in the 2023 OSCAR analysis, limiting that evaluation to precision.

Fourth, the study did not include conventional discriminative NLP baselines, such as rule-based systems or encoder-only based classifiers, which would help determine whether generative modeling is necessary for this task.

Fifth, the training targets used a standardized explanation rather than physician-authored reasoning tailored to each example. Consequently, the generated rationales and supporting quotations were not evaluated for completeness, faithfulness, or clinical usefulness and should not be interpreted as validated explanations. Richer supervision or human-feedback methods may improve evidence selection, but these approaches would require separate evaluation.

Sixth, the fine-tuned Qwen3-4B’s substantial underperformance relative to Llama-405B with self-correction on the OSCAR corpus (line 4) was initially surprising. Our analysis revealed that the fine-tuned Qwen3-4B was largely unable to auto-correct its answers: it almost always confirmed its original decision, and in the rare cases where it did revise its output, the change was limited to the location of the cardiac arrest rather than the underlying classification. This resulted in a large number of false positives that persisted despite the self-correction step—errors that Llama-405B, by contrast, was able to correctly resolve during its own self-correction. We hypothesize that this reflects a loss of calibration following task-specific finetuning: as the model became increasingly specialized in detecting IHCA within the training distribution, it also grew overconfident in its predictions, which is consistent with catastrophic forgetting of the general self-critique behavior present in the base instruction-tuned model. To address this, we are currently testing an alternative pipeline in which Llama-405B performs the correction step on the fine-tuned model’s outputs, rather than the fine-tuned model correcting itself. We expect this to improve overall performance, since the fine-tuned model alone (line 6) already outperforms Llama-405B without self-correction (line 3) with 0.59 vs. 0.47^a^.

Finally, the present results reflect retrospective model development. Prospective implementation would require secure data handling, access controls, audit trails, monitoring for performance drift, and clearly defined clinician responsibility. The model should be used to support, rather than replace, physician adjudication.

## 5. Conclusion

Explicit integration of physician-defined clinical knowledge through guideline-enriched prompting, supervised fine-tuning, and targeted self-correction enabled accurate identification of IHCA events and their locations from EHR notes. A fully fine-tuned 4-billion-parameter model achieved the strongest performance on the curated Evaluation Corpus and performed at least comparably to a substantially larger prompted model.

These findings support the use of locally deployable LLMs as screening tools for retrospective IHCA cohort construction. In a clinician-supervised workflow, the model could prioritize likely IHCA encounters for adjudication and reduce reliance on diagnosis and procedure codes alone. Validation across independent health systems, complete assessment in lower-prevalence cohorts, evaluation of model-provided evidence, and prospective measurement of abstraction efficiency are required before operational deployment or broad registry use.

## Data Availability

Prompt and code produced in the present study are available upon reasonable request to the authors.

## Data Availability

Prompts are available upon reasonable request.

## Code Availability

Code available upon reasonable request.

## Footnotes

a Upon acceptance of the paper, we will report the updated performance.

